# Integrated One Health Surveillance of Zoonotic Diseases at the Wildlife-Human Interface: How Pathogen Detection Informs Spillover Risks and Transmission Dynamics

**DOI:** 10.1101/2025.04.21.25326139

**Authors:** Rajindra Napit, Binod Khadka, Bivisha Khadka, Roji Raut, Pei Yee Woh, Sheina Macy Manalo, Ajit Poudel, Shova Bhandari, Bindiya Bajracharya, Pranav Pandit, Terra Kelly, Jessica S. Schwind, Rajesh Rajbhandari, Bishwo Shrestha, Smriti Khadka, Kavya Dhital, Prajwol Manandhar, Saman Pradhan, Dibesh Karmacharya

**Affiliations:** School of Medicine, Deakin University, Victoria, Australia; One Health Division, Center for Molecular Dynamics Nepal, Kathmandu, Nepal; Department of Food Science and Nutrition, The Hong Kong Polytechnic University, Hong Kong Special Administrative Region; Department of Veterinary Paraclinical Sciences, University of the Philippines Los Banos, The Philippines; EpiCenter for Disease Dynamics, University of California Davis, USA; EpiEcos, USA; Institute for Health Logistics & Analytics, Georgia Southern University, Statesboro, GA, USA; Health Program, Wildlife Conservation Society, New York, USA; School of the Environment, Faculty of Science, University of Queensland, Australia

## Abstract

Emerging infectious diseases (EIDs) pose a significant global health challenge, with zoonotic pathogens— transmitted from animals to humans—accounting for about 75% of these threats. Traditionally, responses to EIDs have been reactive, focusing on post-outbreak containment, which often results in considerable health and economic losses. Recognizing the need for a proactive One Health approach, our study in the buffer zone (Thakurdwara) of Bardia National Park, Nepal aimed to establish a zoonotic disease surveillance program. We conducted disease risk assessments and collected biological samples from 100 households located in the wildlife-human interface of the park. This included one human sample (n=100) and at least two livestock samples per household (n=289). Fecal samples from the surrounding forest were non-invasively collected and identified using DNA barcoding. All samples were screened for ten target pathogens, including six viruses and four bacteria, using polymerase chain reaction (PCR) and DNA sequencing. By integrating laboratory findings with risk survey data using a One Health approach, we analyzed potential zoonotic pathogen spillover and transmission dynamics to better understand the interconnected factors influencing zoonotic disease risks.

*Campylobacter* emerged as the most common pathogen, detected in 97 human households and 219 livestock samples. *Mycobacteria* were identified in two livestock and two wildlife samples, while *Salmonella* was confined to a single human household. Influenza A virus was observed in one livestock sample, and wildlife samples showed low pathogen prevalence overall. *Campylobacter* hotspots were identified through strain variation and network analysis, linking humans, livestock, and poultry, based on proximity, shared water sources, and contamination. Poultry likely served as a reservoir for human cases indicating zoonotic transmission pathways.

Our findings highlight the critical need for integrated One Health measures and proactive, integrated surveillance systems that emphasize early detection, community education, and targeted interventions to mitigate emerging zoonotic threats.

## Background

Emerging infectious diseases (EIDs) represent a significant challenge to global health, affecting not only human populations but also animals and economies. A substantial proportion of these EIDs are zoonotic pathogens, which are diseases that can be transmitted from animals to humans. Research indicates that approximately 75% of newly emerging infectious diseases are zoonotic in origin, highlighting the critical role that wildlife plays in the emergence of these pathogens [1,2]. Among the various types of pathogens, viruses account for a significant proportion of zoonotic diseases, underscoring the importance of understanding viral dynamics in the context of EIDs [3].

Historically, efforts to control zoonotic viruses such as SARS and MERS have been largely reactive, focusing on containment after outbreaks have occurred. This reactive approach often leads to significant loss of life and economic damage, as seen during the SARS outbreak in 2003 and the ongoing challenges posed by COVID-19 [3]. The reactive nature of these responses has prompted a shift towards proactive measures, largely influenced by initiatives such as USAID’s Emerging Pandemic Threats Program. Programs like PREDICT emphasized early identification of emerging diseases at their source, which can be crucial for developing intervention strategies to prevent spillover events before they escalate into widespread outbreaks [2].

Nepal, with its rich biodiversity and extensive wildlife-human interface, exemplifies a region at heightened risk for zoonotic disease outbreaks. This susceptibility is further compounded by cultural practices, ethnic diversity, and traditional beliefs that influence human-animal interactions [4]. Given the critical importance of identifying potential wildlife carriers of zoonotic diseases to mitigate their spread to humans, Nepal’s approach to biodiversity conservation and human-wildlife coexistence is of particular relevance. The country’s establishment of protected areas, including twelve National Parks and various conservation areas, embodies its commitment to preserving biodiversity. However, these conservation efforts also create buffer zones where wildlife and human communities intersect, potentially facilitating the transmission of zoonotic diseases [4,5].

The role of pathogens, including virus discovery, is paramount in preparing for future epidemics. By identifying pathogens in wildlife populations prior to their emergence in humans, researchers can gain insights into the ecology, evolution, and transmission risks associated with potentially pandemic strains[3,6]. For instance, ongoing studies on RNA viruses, particularly coronaviruses in regions like Nepal, aim to elucidate the dynamics of viral spillover and identify potential reservoirs [3]. This proactive surveillance is essential, as it allows for a better understanding of the factors that contribute to zoonotic spillover, including human behaviors and environmental changes that increase the likelihood of pathogen transmission [7].

In addition to identifying pathogens, establishing surveillance systems in areas where environmental and market changes occur is critical for mitigating zoonotic spillover risks. By raising awareness and building capacity among stakeholders, such initiatives can enhance the effectiveness of zoonotic disease surveillance and response strategies [8]. Increasing awareness of zoonotic diseases and the need for proactive surveillance systems are vital in addressing the challenges posed by emerging infectious diseases. Understanding the mechanisms of pathogen spillover and the dynamics of host-pathogen interactions will be crucial for developing effective strategies to mitigate the risks associated with zoonotic pathogens.

Our study aimed to establish an active zoonotic disease surveillance and mitigation program, starting with a pilot study in Bardia National Park and its surrounding buffer zone. It sought to gather baseline data on zoonotic diseases in nearby communities, raise awareness of proactive surveillance systems, and collect biological samples for molecular analysis. By collecting and analyzing blood, oral, and stool samples from humans, livestock, and wildlife, the study identified pathogens present, assessed the risks of zoonotic spillover, and evaluated the potential for disease transmission between humans and wildlife. This comprehensive approach also focused on identifying risky local behaviors, building capacity through stakeholder training in sample collection and laboratory processing, and informing public health strategies and interventions in Nepal and similar contexts worldwide, where biodiversity and human populations closely interact.

## Methodology

### Site and Sampling

Bardia National Park (28.3833° N and 81.3333° E), one of Nepal’s largest national parks with surrounding buffer zone, is home to over 130,000 people across several municipalities in Bardia, Banke, and Surkhet. These communities rely on wildlife interactions for their livelihoods, including eco-tourism. The park, rich in biodiversity with species like Bengal tigers, Indian elephants, gharials, and one-horned rhinoceroses, also attracts national and international tourists, further elevating transmission risks. Thakurdwara Municipality, adjacent to the park, has 8,772 residents, primarily Tharu, along with Brahmin, Chettri, and Dalit groups. Livestock farming dominates the local economy, while a growing tiger population has heightened human-wildlife conflicts. Buffer zone communities depend on park resources like grass, fodder, and medicinal herbs, intensifying human-wildlife interactions. These dynamics, combined with tourism, highlight the need for sustainable management practices to mitigate zoonotic risks and foster coexistence.

### Survey Procedures

#### One Health Risk Assessment Survey

One-Health risk assessment survey of all households was conducted using a standardized questionnaire, assessing factors that contribute to risks associated with transmission of zoonotic diseases between human, livestock and wildlife interfaces from 5th July 2023 to 26th July 2023. The survey questionnaire also captured other important disease risk factors such as water quality, sanitation, and animal handling practices. Collected data were digitized, cleaned, and statistical analysis was conducted using IBM SPSS 20.

Enumerators were trained to administer survey questionnaires following the Center for Molecular Dynamics Nepal (CMDN) sampling strategies-ensuring representative and unbiased sample collection.

#### Engagement with Local Stakeholders

All pertinent stakeholders (local authorities, government and national park officials, and community leaders) were informed of the study activities in a workshop to ensure their cooperation. Field staff, including two local phlebotomists and veterinary technicians, were trained by the CMDN on biosafety and biosecurity practices, maintenance of cold chain and proper waste management. to build local capacity and support sampling efforts.

Ward Number 9 of Thakurdwara was selected for the study given its proximity to the national park, and data was collected on human and livestock from 100 households (September-November 2023) (Figure 2). Data was first gathered from the local Ward Offices and a screening survey was conducted to identify households with livestock, excluding those without. Simple random sampling (SRS) was then used to select households for inclusion in the study.

**Figure 1:**
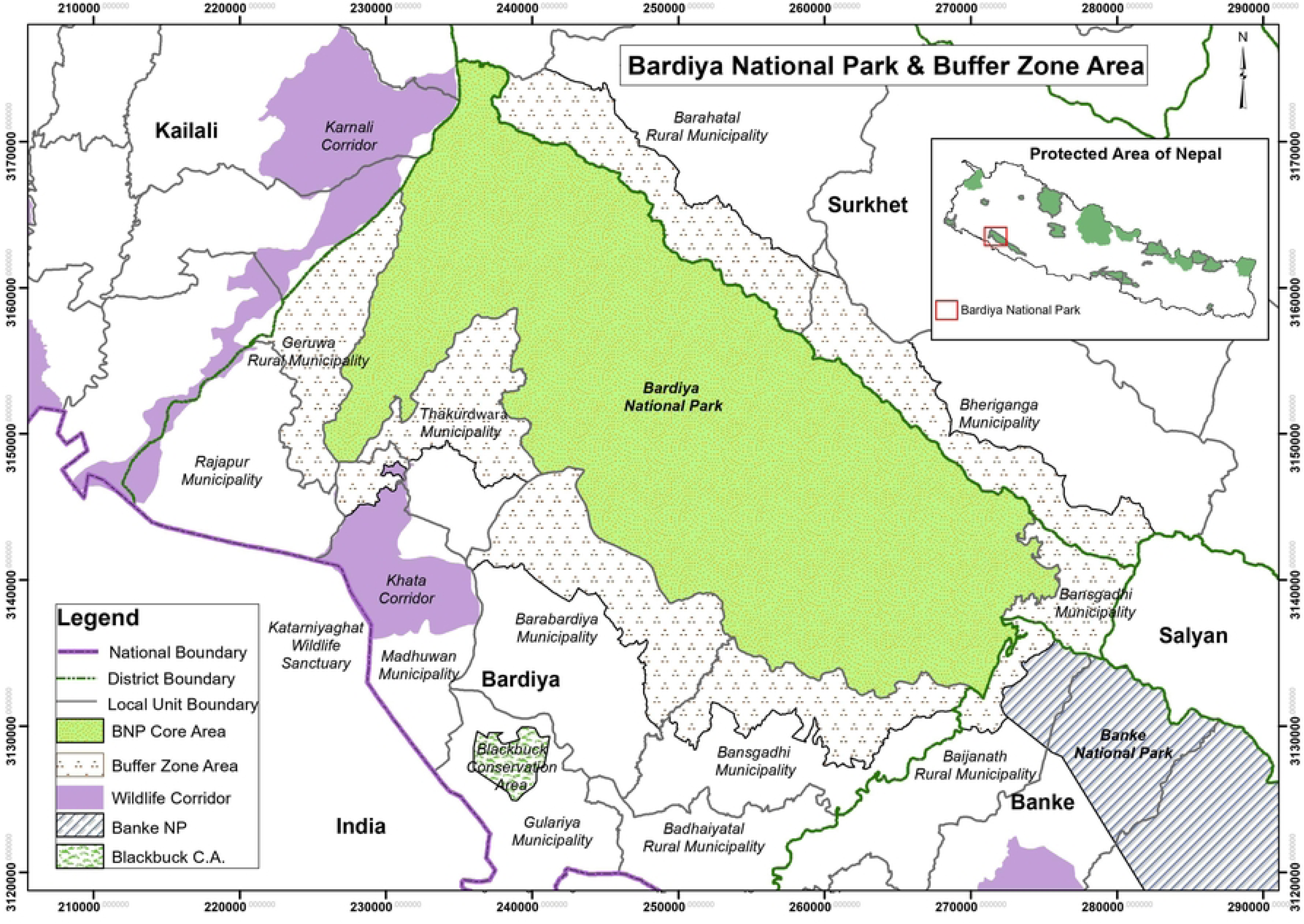
Location of Thakurdwara Community in the buffer zone of Bardia National Park *(source:* https://bardianationalpark.gov.np/en/others/1/)

**Figure 2.**
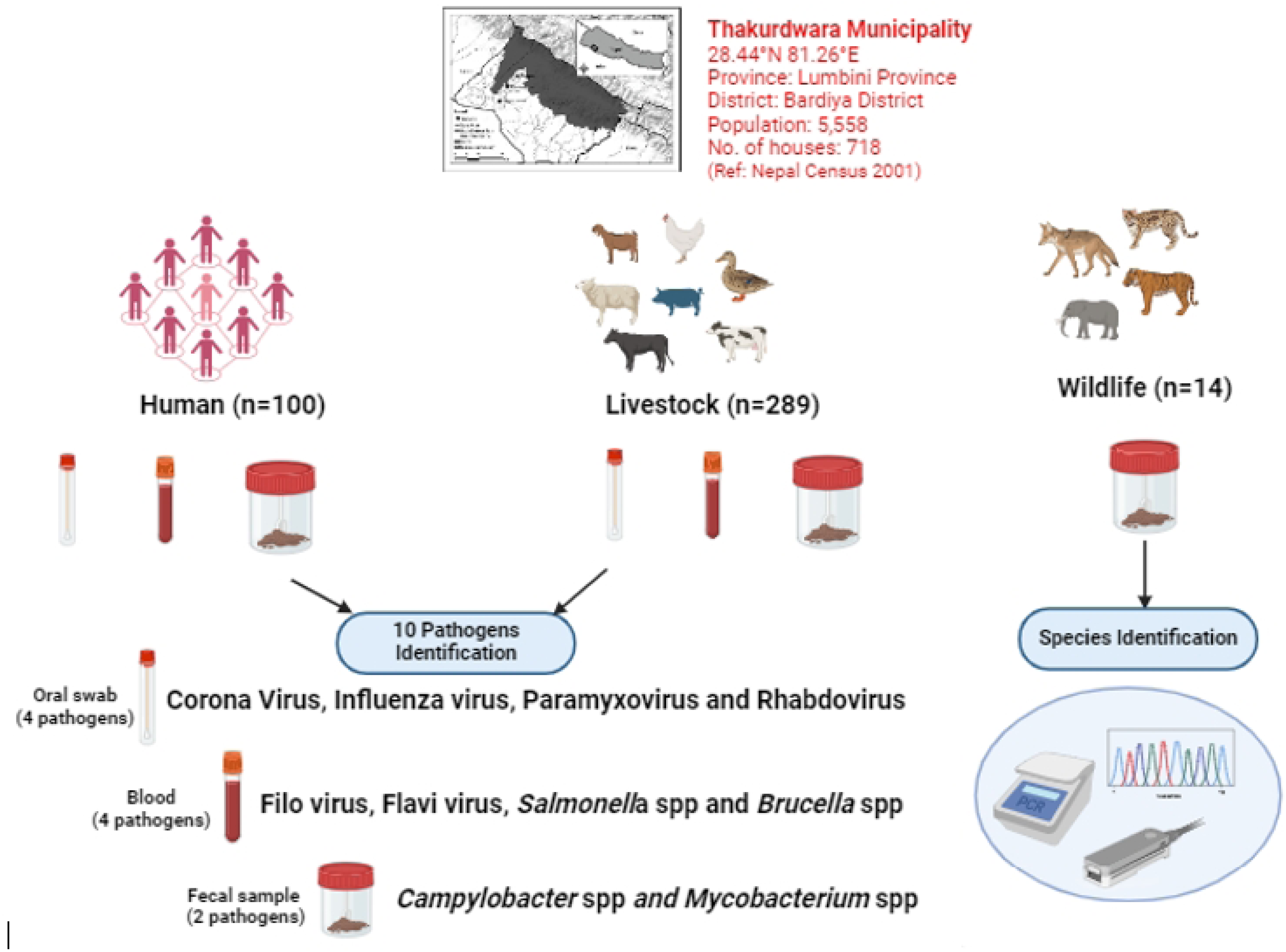
Sample collection and pathogen screening for human, livestock, and wildlife samples from Thakudwara-9, Bardia National Park, conducted at the CMDN Lab. The figure was created with Biorender [9].

#### Ethical Consideration

Written consent was obtained from all participants in the study to collect samples from them and their livestock. Trained phlebotomists and veterinary technicians were used to collect samples from humans and livestock respectively. Livestock were sampled in the presence of their primary caretaker. Ethical guidelines of the Department of Livestock Services (DLS) were followed, and ethical approval was obtained from the Nepal Health Research Council (Registration Number – 272/2023).

### Pathogen Selection and Screening

We chose ten common pathogens for the study based on their endemicity and the availability of detection assays for screening (Supplementary Table 2). These included six RNA viruses (*Influenza A*, *Rhabdovirus*, *Paramyxovirus*, *Filovirus*, *Flavivirus* and *Coronavirus*) and four bacteria (*Salmonella*, *Brucella*, *Campylobacter* and *Mycobacteria*). The justification for pathogen selection, along with their zoonotic potential and transmission pathways, is provided in the Supplementary Table 1.

### Sample Collection and Processing

The study involved the collection of various sample types, including blood in EDTA, blood in viral transport media (VTM), oral swabs, stool in PBS, and wildlife scat samples. A total of 100 humans, 289 livestock, and 14 wildlife fecal samples were collected. For each human and livestock subject, three types of samples—blood, oral swab, and stool—were taken. Trained personnel collected oral swabs from humans and livestock using labeled cryotubes containing 2 ml of VTM. Blood samples were drawn through venipuncture into EDTA tubes, and stool samples were placed in labeled, sterile, wide-mouthed containers and suspended in PBS. Wildlife fecal samples were collected in sterile falcon tubes. All samples were transported to the laboratory under cold chain conditions, maintaining a temperature of 2-8°C. Upon arrival at the lab, oral swabs in VTM, blood in VTM, and stool samples in PBS were stored at -20°C, while blood in EDTA were stored at 4°C (Figure 2).

Nucleic acid extraction for oral swabs in VTM and blood in EDTA was performed using the abGeneix Automated DNA and RNA extraction system (REF 800800) and Uni-medica nucleic acid extraction reagent, following the manufacturer’s instructions. Stool samples in PBS and wildlife fecal samples were extracted using the QIAamp Fast DNA Stool Mini kit using manufacturer’s instruction.

cDNA was synthesized using the BIO-RAD iScript cDNA synthesis kit. The reaction was carried out in a 20 µl volume containing 4 µl of 5X iScript reaction mix, 1 µl iScript reverse transcriptase, 6 µl NFW, and 9 µl template RNA. Both RNA virus and bacterial pathogen detection were conducted using PCR protocols referenced in the Supplementary Table 2.

To confirm DNA and RNA extraction, a quality control PCR was performed on all oral, blood, and stool samples, targeting the universal mammal CytB gene (457 bp amplicon) using the primers CytB_F (5′-GAGGMCAAATATCATTCTGAGG-3′) and CytB_R (5′-TAGGGCVAGGACTCCTCCTAGT-3′). The PCR was carried out in a 25 µl volume with 12.5 µl Qiagen Mastermix, 1.5 µl each of the primers (10 pmol), 7.5 µl NFW, and 2 µl template DNA. The amplification conditions were initial denaturation at 95°C for 15 minutes, followed by 50 cycles of 94°C for 30 seconds, 52°C for 50 seconds, and 72°C for 60 seconds, with a final extension at 72°C for 7 minutes. PCR products were visualized on a 1.5% gel using a 100 bp DNA ladder.

### Species Identification (DNA barcoding) of Wildlife Fecal Samples

Opportunistic fecal samples (n=14) from the surrounding forest area were collected, and DNA was extracted using the QIAamp Fast DNA Stool Mini kit. PCR was performed in a 25 µl volume with 12.5 µl 2X Amplitaq, 1.5 µl each of CytB_F and CytB_R primers (10 pmol), 0.5 µl NFW, and 5 µl template DNA targeting Cytochrome b of mitochondrial DNA. Amplification was done in a ProFlex thermocycler with initial denaturation at 95°C for 10 minutes, followed by 40 cycles of 95°C for 30 seconds, 50°C for 30 seconds, and 72°C for 60 seconds, with a final extension at 72°C for 10 minutes. PCR products were visualized on a 1.5% gel using a 100 bp DNA ladder, and positive DNA amplicons were further sequenced and species identified using NCBI reference database.

### Characterization of Detected Pathogens Using Nanopore Sequencing Platform

Following PCR analysis, a substantial number of samples were presumptively positive for pathogens including *Campylobacter*, Rhabdovirus, Paramyxovirus, *Brucella*, *Salmonella*, Influenza, and Filoviru*s* in PCR screening. To confirm these results, we performed molecular characterization and validation using next-generation DNA sequencing on PCR amplicons.

The sequencing was carried out on an Oxford Nanopore MinION (MK1C) device. The sequencing was carried out with a dual barcoding approach with the SQK-NBD114.24 kit and EXP-PBC001 on an R10.4.1 flow cell. Sample quantification was carried out using the Qubit HS dsDNA assay kit, while size estimation was assessed through agarose gel electrophoresis.

### Characterization of *Campylobacter* with Phylogenetic Analysis

The raw sequences from Nanopore sequencer were first assessed for quality with Nanoplot v1.42.0 [10] followed by adapter sequences trimming with Porechop v 0.2.4 [11]. The quality filtering was performed with Filtlong v0.2.1 [12] with minimum length and Q-score of 300bp and seven respectively. The cleaned sequence reads were then subjected to variant calling and consensus were generated with Medakka v2.1 [13]. Thus, generated consensus sequences were then used to develop a phylogenetic tree reconstructed from an ∼862 bp alignment of 417 16S rRNA gene sequences representing major species of *Campylobacter*. The tree was inferred using the Maximum Likelihood method in IQ-TREE v1.6.12 [14], applying the best-fit model parameters (GTR+F+I+G4) determined by the Akaike Information Criterion (AIC) in ModelFinder. Node values represent bootstrap support scores (1,000 replicates) from the Maximum Likelihood test. Reference sequences, denoted by black tips, were retrieved from GenBank, and their taxonomy was inferred from corresponding publications. Sequences generated in this study using MinION amplicon sequencing are indicated by red tips (human-derived) and blue tips (livestock-derived). Lineages or cluster groups corresponding to known *Campylobacter* species were annotated with abbreviations. The 16S rRNA gene sequence of *Arcobacter butzleri* (GenBank accession: JQ743025) was used as an outgroup to root the *Campylobacter* species tree.

### Network Analysis of the most Prevalent Pathogen-*Campylobacter*

The presence of *Campylobacter* in almost all households and different livestock and humans were used to perform a network analysis. Incident data (human and animals) along with sub-species level classification data obtained from 16S *Campylobacter* phylogeny was utilized to create a matrix table. The table was then imported into Gephi v0.100 to create the network diagram using default settings. The stats were calculated with the built-in analysis plugin. The different communities were identified using the modularity function (using node weight, resolution of 1, and randomization ‘on’) within Gephi v0.10.

## Results

### Demographic and Socio-economic Profile of the Community

Thakurdwara is a rural community with a median population of 36 years. Most respondents were married or widowed (86%), and a significant portion had a limited education, with 31% being illiterate and 26% having completed higher secondary school. Agriculture was the primary occupation, with 83% of individuals engaged in farming, while smaller groups worked in skilled (4%) and unskilled manual labor (2%). Households typically raised poultry (94%), goats (82%), and cattle (68%), with swine (46%) and dogs (22%) also commonly kept. Medical care was mainly sought at clinics or health centers (94%), with fewer people visiting hospitals (48%) or consulting community health workers (4%). Over the past year, common symptoms included fever with muscle aches (15%), while cases of encephalitis (7%) and hemorrhagic fever (1%) were less frequent. Most households depended on uncovered wells, ponds, or rivers for water (92%), with only half (49%) filtering it before use. Additionally, 72% of residents shared water for animal feeding (Table 1).

**Table 1.** Demographic, medical history, and drinking water condition of respondents (n=100)

| **Characteristics** | **Frequency**<br>**%(n)** |
| --- | --- |
| ***Demographic*** |  |
| **Age** (median [IQR]) | 36 [16-84] |
| **Marital status** |  |
| Unmarried | 14 |
| Married/widowed | 86 |
| **Educational level** |  |
| Illiterate | 31 |
| Primary (Grade 1-5) | 17 |
| Secondary (Grade 6-8) | 10 |
| Higher Secondary (Grade 9-10) | 26 |
| Intermediate | 12 |
| Diploma | 4 |
| **Occupation** |  |
| Professional/technical/managerial | 7 |
| Skilled manual | 4 |
| Unskilled manual | 2 |
| Agriculture | 83 |
| Meat production | 1 |
| Others | 5 |
| **Household livestock/animal raised** |  |
| Poultry | 94 |
| Mallard | 12 |
| Cattle | 68 |
| Goat | 82 |
| Sheep | 4 |
| Swine | 46 |
| Dog | 22 |
| Cat | 12 |
| Others | 2 |
| ***Medical record*** |  |
| Main medical facility to receive medical treatment |  |
| Clinic/health center | 94 |
| Hospital | 48 |
| Community health worker | 4 |
| Dispensary/pharmacy | 1 |
| Others | 3 |
| Ever had an unusual fever during the past 12 months |  |
| Fever with headache and severe fatigue or weakness (encephalitis) | 7 |
| Fever with bleeding or bruising not related to injury (hemorrhagic fever) | 1 |
| Fever with cough and shortness of breath or difficulty breathing | 5 |
| (Severe Acute Respiratory Infections (SARI) or COVID-19) |  |
| Fever with muscle aches, cough, or sore throat | 15 |
| Fever with diarrhea or vomiting | 4 |
| Persistent rash or soreness on skin | 1 |
| None of these symptoms | 9 |
| No fever | 65 |
| ***Usage of drinking water*** |  |
| Source of water |  |
| Piped-in water/water taps | 9 |
| Uncovered well/pond/river | 92 |
| Treatment of water |  |
| Filtration | 49 |
| No treatment | 51 |
| Shared water for humans and animals |  |
| Yes | 72 |
| No | 28 |

### Living Conditions, Basic Amenities and Health Hygiene Practices

Nearly half of the households surveyed (48%) had permanent housing structures, while the rest lived in more vulnerable, temporary dwellings. In terms of sanitation, 20% of households lacked any toilet facilities, with the majority (56%) using pit latrines with smaller portions using compost toilets (12%) or flush toilets (12%). Access to clean water was limited, with only 8% households having piped water, while the vast majority (92%) relied on uncovered wells, ponds, or rivers. Of the 100 people surveyed, 35% reported having been ill in the past year. Among them, 18% did not know the cause of their illness, 7% attributed it to contaminated food or water, and 5% believed they contracted the disease from other infected individuals.

#### Animal Handling Practices

Nearly all respondents handled and raised live animals with the majority (78%) sharing water sources with animals. High risk exposure activities were also reported with 26% of respondents mentioning slaughtering animals, 34% eating food touched by animals, and 20% consuming raw or undercooked meat (Table 2). Meanwhile, 45% reported eating food contaminated with animal feces.

**Table 2.** Animal handling practices amongst the respondents (n=100).

| Characteristics | Frequency<br>% |
| --- | --- |
| Consumed food touched or damaged by animals |  |
| Yes | 35 |
| No | 65 |
| Allowed animals to come inside the living place |  |
| Yes | 56 |
| No | 44 |
| Presence of sick animals |  |
| Yes | 13 |
| No | 87 |
| Handled meat/organs/blood of killed animals |  |
| Yes | 59 |
| No | 41 |
| Eaten raw or undercooked meat/organs/blood |  |
| Yes | 20 |
| No | 80 |
| Eaten unwell or sick animals |  |
| Yes | 21 |
| No | 79 |
| Eaten or shared collected dead animals |  |
| Yes | 14 |
| No | 86 |
| Sold deadstock or fallen stock animals |  |
| Yes | 5 |
| No | 95 |
| Scratched or bitten by animals |  |
| Yes | 6 |
| No | 94 |
| Slaughtered an animal |  |
| Yes | 26 |
| No | 74 |
| Hunted or trapped an animal |  |
| Yes | 2 |
| No | 98 |

#### Wildlife-Livestock Interactions

Wildlife-livestock interactions were reported by 31% of respondents, mainly involving tigers (35.5%), elephants (25.8%), and leopards (19.4%). Direct encounters with wildlife were also reported, including with rhinos (33%), tigers (29%), and elephants (29%).

### Pathogen Detected in Human, Livestock and Wildlife Samples

Among human samples, *Salmonella* was detected in one case, while *Campylobacter* was found in 97 households, making it the most common human pathogen (Table 3). In livestock, Influenza A virus was identified in one livestock sample, while *Campylobacter* was widespread, appearing in 219 samples. *Mycobacteria* were detected in two livestock samples.

**Table 3:** Pathogens detected in the study from n=403 samples.

|  |  | Pathogen | Detected in Samples |
| --- | --- | --- | --- |
| Humans<br>(n=100) | Bacteria | *Salmonella* | 1 |
|  |  | *Campylobacter* | 97 |
| Livestock<br>(n=289) | Virus | Influenza A<br>Virus | 1 |
|  | Bacteria | *Campylobacter* | 219 |
|  |  | *Mycobacteria* | 2 |
| Wildlife<br>(n=14) | Bacteria | *Campylobacter* | 2 |
|  |  | *Mycobacteria* | 2 |

Using DNA barcoding, we identified fecal samples from two wild species, the Asiatic elephant (*Elephas maximus*) and hog deer (*Axis porcinus*)—in the forest surrounding Thakurdwara. These samples showed low pathogen prevalence, with *Campylobacter* and *Mycobacteria* each found in two cases. Overall***, Campylobacter*** was the most commonly found pathogen across livestock and humans. *Mycobacteria* appeared at much lower frequencies in both livestock and wildlife. **Figure 3** illustrates various pathogens detected across hosts (human, livestock including poultry, and wildlife) in the community.

**Figure 3.**
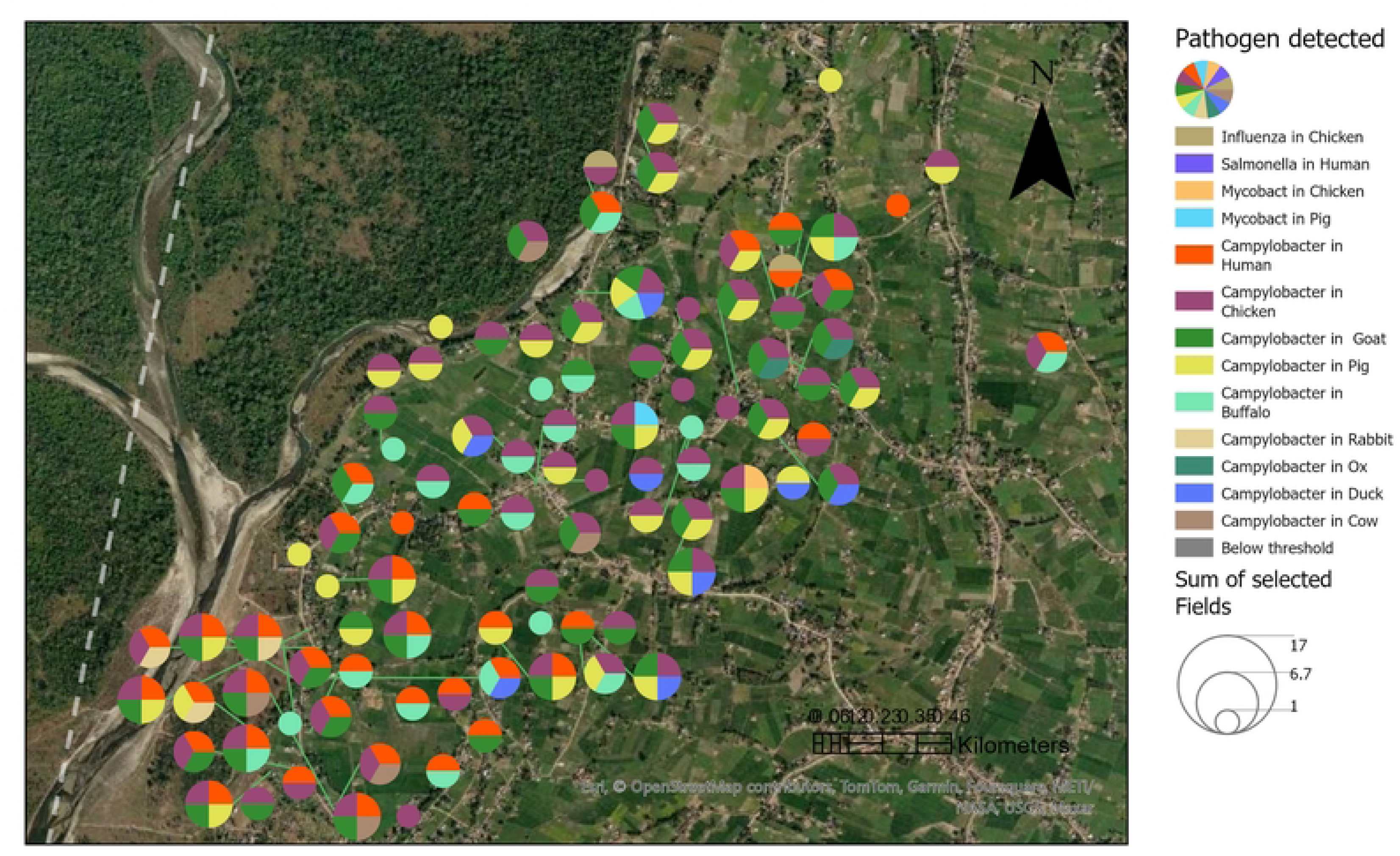
**Spatial map visualizing pathogen distribution across the study site, with pie charts representing various detected pathogens in humans and animals.** Pathogens include *Influenza A*, *Salmonella*, *Mycobacterium*, and multiple *Campylobacter* species, with colors indicating specific hosts. The size of each chart reflects pathogen detection frequency, with the highest numbers of detections in the lower left and central regions, suggesting zoonotic disease hotspots near settlements and water bodies.

#### Associations between Animal Handling Practices and Detected Pathogens in Humans

*Campylobacter* was detected commonly detected pathogen in high-risk animal contact scenarios. Handling and raising live animals had the highest *Campylobacter* detections (97 and 96 cases, respectively), along with a few *Salmonella* and *Mycobacteria* cases. Sharing water with animals for washing was linked to 76 *Campylobacter* and one *Salmonella* detection. Other risky practices, such as eating near animal feces (44 *Campylobacter*), consuming food touched by animals (33 *Campylobacter*), and keeping animals inside homes (53 *Campylobacter*), also were found. Cooking or handling recently killed animals resulted in 58 *Campylobacter*, one *Salmonella*, and one *Mycobacteria* detection.

#### Distribution of *Campylobacter Strains* across Diverse Species within the Community

We created a geo-spatial map to visualize all samples with detected *Campylobacter* (Figure 4) and analyzed detected strains (Supplementary Table 3, Figure 5) using a phylogenetic tree (Figure 5). Our analysis revealed distinct *Campylobacter* clusters that were genetically identical in both humans and animals.

**Figure 4:**
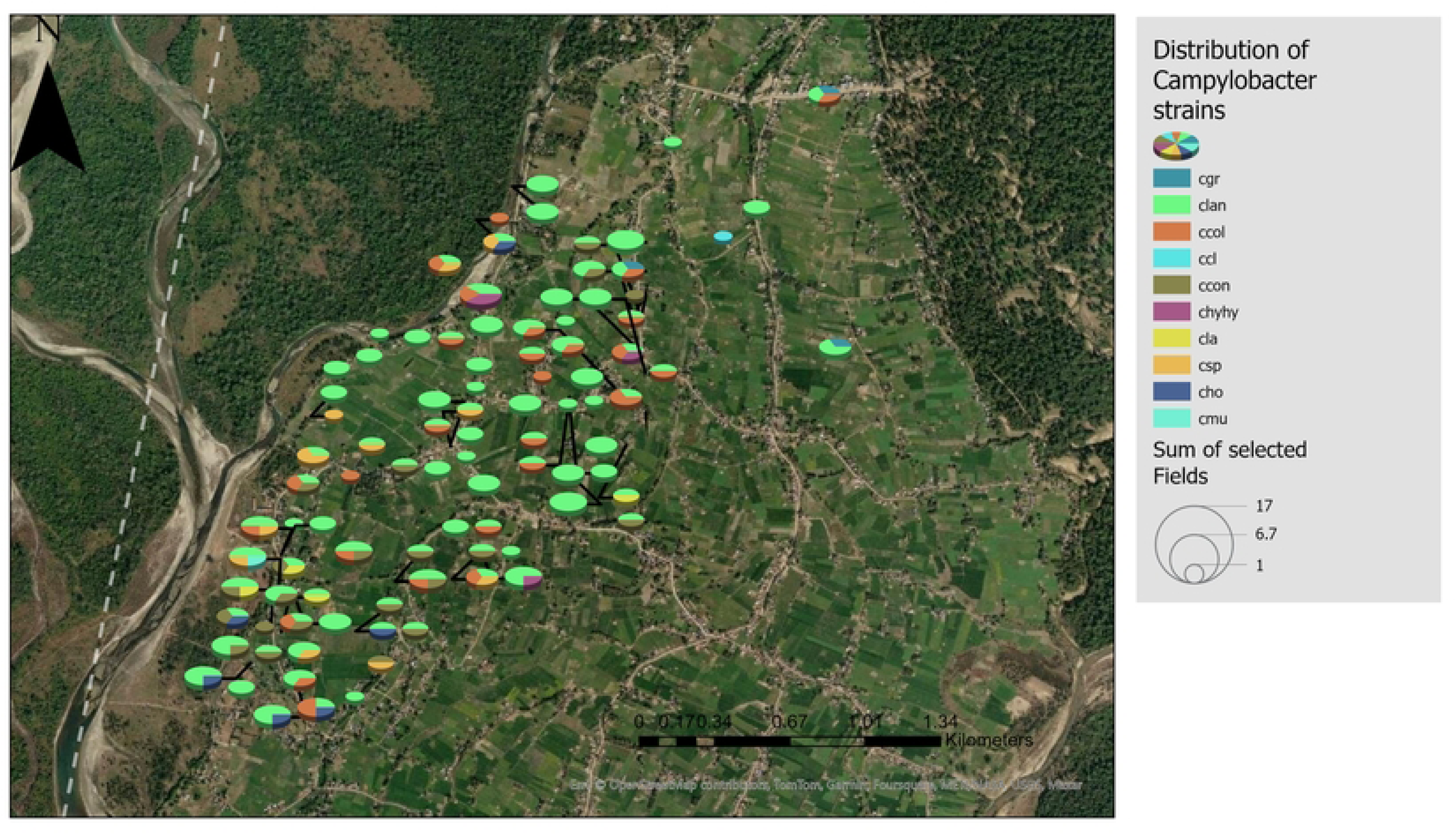
**Spatial distribution of *Campylobacter* in a community**, with detection points in humans, livestock, and poultry. Clusters of overlapping cases indicate zoonotic hotspots where transmission is likely amplified by proximity, shared water sources, or environmental contamination, while outlier cases suggest sporadic exposure events. Poultry appears to be a potential reservoir, with zoonotic transmission risks. The map underscores the interconnected role of humans, livestock, and poultry in bacterial spread.

**Figure 5:**
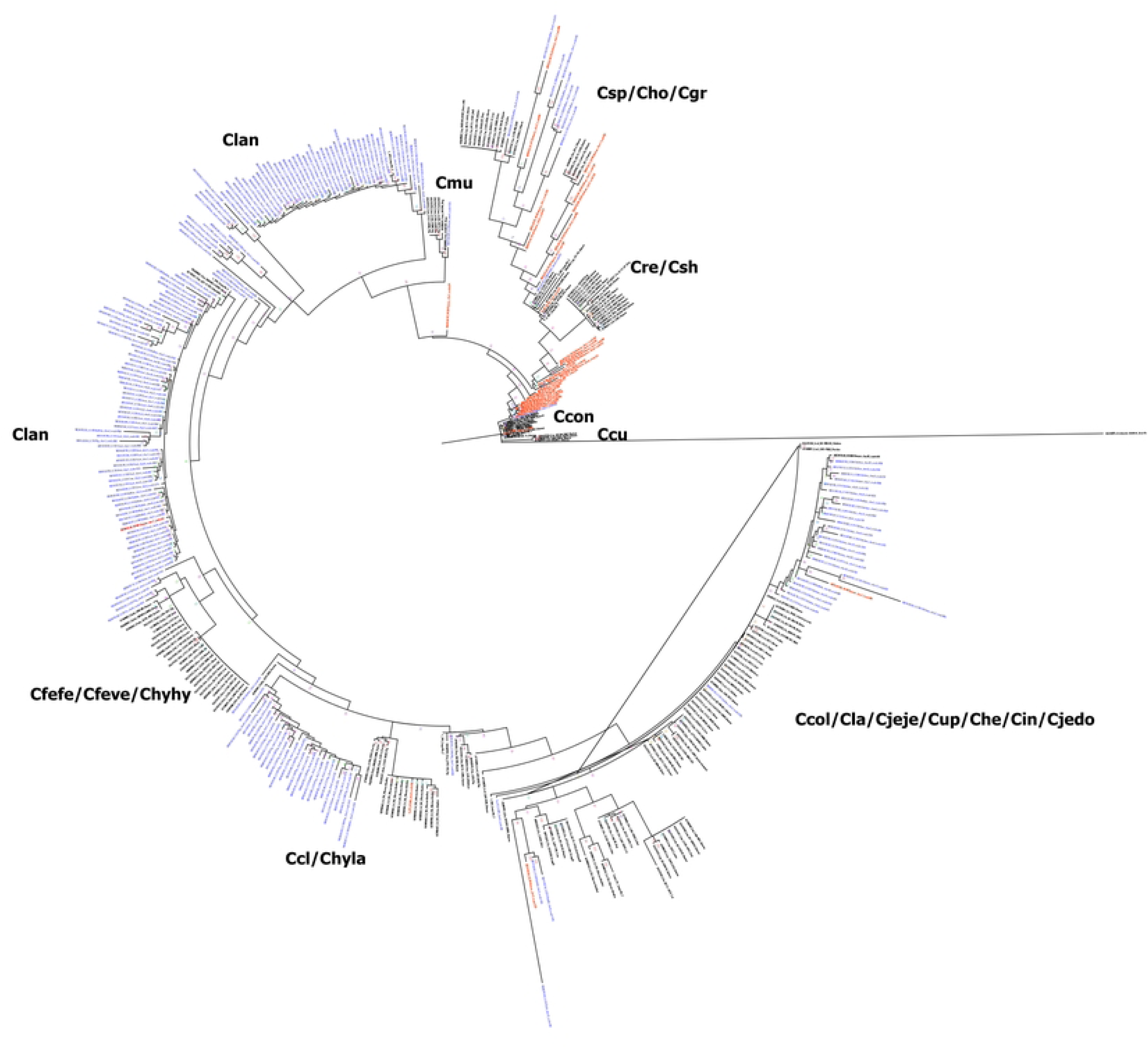
**Phylogenetic analysis of Campylobacter detected in fecal samples from humans and livestock in the Thakurdwara buffer zone of Bardia National Park,** 2023. Phylogenetic tree reconstructed from an ∼862 bp alignment of 417 16S rRNA gene sequences representing major species of *Campylobacter*. Sequences generated in this study using MinION amplicon sequencing are indicated by red tips (**human-derived**) and blue tips **(livestock-derived**). Lineages or cluster groups corresponding to known *Campylobacter* species are annotated with abbreviations, as follows: *Ccol:* C. coli*, Cla:* C. lari*, Cjeje:* C. jejuni subsp. jejuni*, Cup:* C. upsaliensis*, Che:* C. helveticus*, Cin:* C. insulaenigrae*, Cjedo:* C. jejuni *subsp.* doylei*, Ccl:* C. species *clone, Chyla:* C. hyointestinalis *subsp.* lawsonii*, Cfefe:* C. fetus *subsp.* fetus*, Cfeve:* C. fetus *subsp.* venerealis*, Chyhy:* C. hyointestinalis *subsp.* hyointestinalis*, Cmu:* C. mucosalis*, Csp:* C. sputorum*, Cho:* C. hominis*, Cgr:* C. gracilis*, Cre:* C. rectus*, Csh:* C. showae*, Ccon:* C. concisus*, Ccu:* C. curvus; *Clan*:*C. lanienae.* The 16S rRNA gene sequence of *Arcobacter butzleri* (GenBank accession: JQ743025) was used as an outgroup to root the *Campylobacter* species tree.

In one of the *Campylobacter* clusters, the samples included a duck, two rabbits, and a human, all sharing the same *Campylobacter* strain and located in close geographic proximity. Similarly, another cluster of *Campylobacter* comprised samples from a human, buffalo, and chicken, which were located within a 1.92-kilometer radius.

This phylogenetic tree (Figure 5) illustrates the evolutionary relationships among *Campylobacter* strains, with distinct clusters marked by color-coded annotations. The branching structure highlights genetic variations, with closely related strains grouping together. Labeled clades, such as *Ccol, Cla,* and *Cjeje*, represent different *Campylobacter* species or subspecies, while some clusters contain multiple species, indicating possible genetic overlap. Sample labels, displayed in various colors, may correspond to different host origins, suggesting zoonotic transmission. The tree provides insights into strain diversity and potential cross-species transmission patterns, making it useful for understanding *Campylobacter* epidemiology.

#### Network Analysis Underscoring the Interconnectedness of Livestock, Wildlife, and Humans in Sustaining and Spreading *Campylobacter*

Network analysis based on ∼862 bp alignment of 417 16S rRNA gene sequences representing major species of *Campylobacter* identified a highly connected dense connectivity among livestock (these highly connected nodes representing-goat, chicken, pig, cow, duck, buffalo) (Figure 6). Chickens and pigs appear to be central nodes, suggesting they play a major role in transmission dynamics. The human node is linked to multiple animal hosts, especially chicken and goat. Some nodes (e.g., rabbit, buffalo) connect to multiple species. However, modularity-based analysis separated the networks into five highly related distinct communities based on the interconnectedness of the nodes. The densely connected goat, chicken, pig and ox appears to be the largest community, probably pointing to intricate One Health interactions among these animals with three prominent subspecies of *Campylobacter*. Similarly, humans and rabbits appear to have formed a separate community with subspecies-*Ccl/Ccon* (C. species *clone/* C. *concisus*) and buffalo and cow-both formed separate unique communities with no other animals or humans. However, there was one unique community of households with subspecies of *Cgr* (*C. gracilis*) that did not form a community with any specific animals or humans, indicating the *Campylobacter* spillover and interaction might be more diverse. The network clearly shows the predominant species of *Campylobacter* among different communities of households along with potential One Health interface and interactions. Furthermore, these connections could represent *Campylobacter* presence in shared environments such as water, soil, or food supply chains.

**Figure 6:**
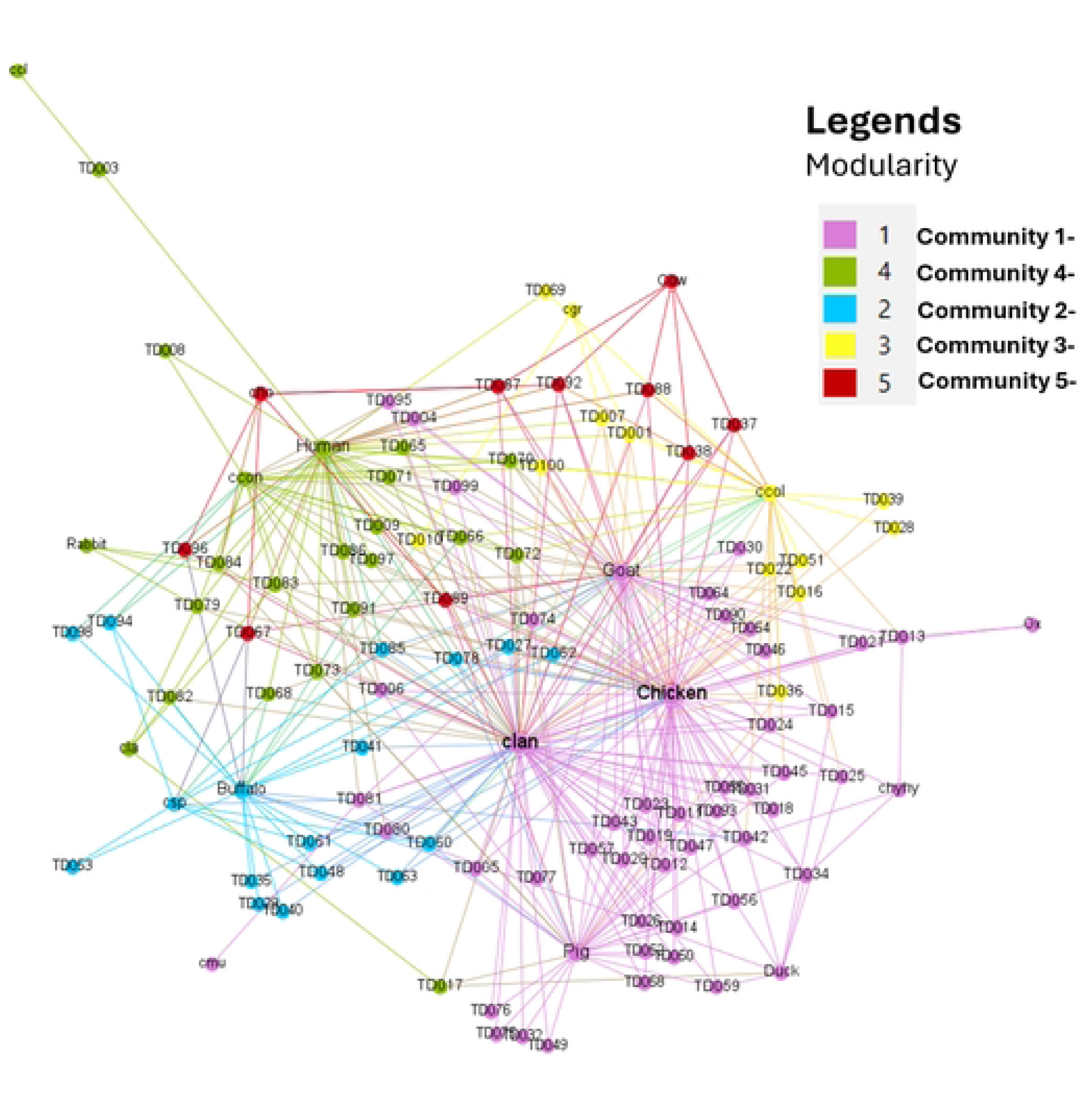
**Network analysis underscoring the interconnectedness of livestock, wildlife, and humans in sustaining and spreading *Campylobacter*. Labeled nodes (e.g., human, goat, chicken, cow, pig, buffalo, rabbit, duck, etc.)** represent host species or environmental reservoirs. **TD codes (e.g., TD001, TD095, TD009, etc.)** represent individual households, potentially linking to genetic sequences or sources of *Campylobacter*.

#### Mycobacterium detection in wildlife and livestock samples

We detected *Mycobacterium* spp. in both wildlife and livestock samples. 16S rRNA gene sequencing and subsequent phylogenetic analysis indicated the opportunistic samples from elephants and hog deer belonged to the *Mycolibacterium* group, while fecal samples from pigs (TD19) and chickens (TD20) clustered within the *Mycobacteroide**s*** group (Figure 7).

**Figure 7:**
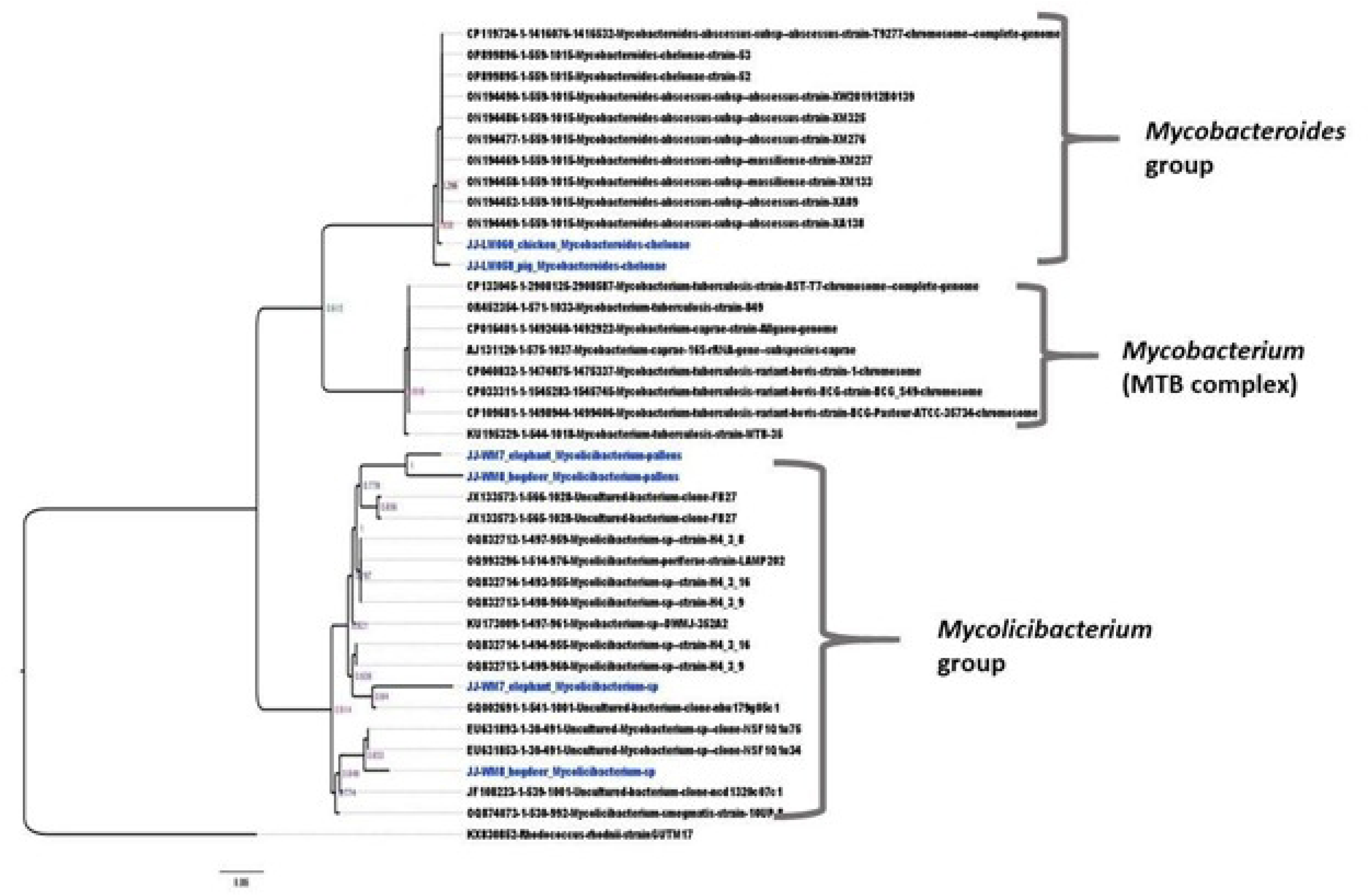
**Phylogenetic analysis of *Mycobacterium*-positive samples, including the *Mycobacteroides* group, MTB complex, and** Mycolibacterium **group**. The *Mycobacterium* (MTB complex) includes pathogenic strains like *M. tuberculosis* and *M. bovis,* with host-specific isolates from pigs and chickens. The ***Mycolicibacterium* group** contains environmental and host-associated strains, including those from elephants and hog deer.

## Discussion

### Public Health Risks and Pathogen Transmission Dynamics in Thakurdwara

This study highlights public health risks from human-animal interactions in Thakurdwara, where zoonotic disease transmission is influenced by socio-economic, environmental, and behavioral factors. With a low literacy rate (32%), effective disease prevention remains a challenge. Despite 52% awareness of zoonotic risks, high-risk behaviors persist, including handling live animals (100%), sharing water sources with livestock (78%), and consuming food near animal feces (45%).

Livestock ownership is common, with chickens (30.3%), goats (27.4%), and cattle (23.2%) being the most prevalent. Wildlife-livestock interactions were reported by 31% of participants, mainly involving tigers (35.5%), elephants (25.8%), and leopards (19.4%). Direct encounters with wildlife during open grazing in the buffer zones heighten the risk of cross-species transmission.

These risks are further amplified by poor sanitation and limited access to safe water. About half (48%) of households live in non-permanent structures, 20% lack toilets, and majority (92%) rely on uncovered wells for their water supply. Only 8% have piped water, and few households treat drinking water, increasing their risk of water-borne infections. Similar practices were observed in other studies in Nepal, which were correlated with high prevalence of the waterborne diseases [15].These conditions and interconnected risks continue to be a public health concern, particularly in vulnerable households in Thakurdwara.

### Pathogen Ecology, Reservoirs, Spillover, and Transmission Dynamics

Network analysis revealed that livestock, particularly poultry, act as primary reservoirs for *Campylobacter* spillover, highlighting the need for targeted interventions in poultry farming, slaughterhouse operations, and food hygiene practices to mitigate transmission risks. The identification of distinct household communities based on *Campylobacter* interconnectedness suggests complex transmission dynamics driven by One Health interactions. Humans formed a unique cluster with rabbits, supporting the likelihood of bidirectional disease spillover, as rabbits are commonly used as model organisms for human diseases [16,17].

Based on the network analysis, the largest host community included chickens, goats, pigs, and oxen, while buffalo and cows formed distinct, isolated communities, indicating minimal One Health interactions with these animals. Surprisingly, the second-largest community lacked specific host species but connected with multiple other communities, suggesting that a particular *Campylobacter* subspecies (*C. ccol*) has spilled over to multiple hosts, reflecting a highly dynamic One Health interface. Shared nodes among different *Campylobacter* strains further indicate indirect transmission pathways through contaminated water, feed, or environmental surfaces.

Geospatial analysis of these communities revealed distinct geographical separations with overlapping regions where One Health interactions occur. Community which lacks clear links to specific animals or humans and is dispersed across the map and often found near or between other communities, potentially representing hotspots for zoonotic spillover. Additionally, different *Campylobacter* subspecies clustered with specific host species, suggest host preferences, a phenomenon observed in *C. jejuni*, which exhibits distinct host specificity [18,19]. However, given the high zoonotic potential of *Campylobacter* spp. [20–22], cross-species transmission remains a significant concern.

Past studies have confirmed the high prevalence of *Campylobacter* in humans and livestock. A meta-analysis in Iran found *Campylobacter* in 43.9% of white meat samples, with chicken at 48.6%, red meat at 7.9%, buffalo at 13.5%, and cattle at 8.4%. Contamination was also found in 19.4% of vegetable samples. [23]. A review in Ethiopia found that *Campylobacter* prevalence was higher in animals (14.6%) than humans (9%) and noted significant antimicrobial resistance [24]

In our study human exposure was closely linked to multiple animal hosts which emphasizes the need for integrated disease surveillance and control strategies across veterinary, environmental, and public health sectors. In addition to *Campylobacter*, *Salmonella* was detected in one human sample, while *Influenza A* appeared in one livestock sample. *Mycobacteria* were identified in two livestock and two wildlife samples, with phylogenetic analysis revealing distinct bacterial clusters between pigs, chickens (*Mycobacteroides* group), and elephants, hog deer (*Mycolibacterium* group), indicating diverse transmission sources.

### Key Drivers of Zoonotic Disease Spread

Geo-spatial mapping and network analysis visually demonstrated the interconnections between humans, livestock, and wildlife in pathogen maintenance and transmission. The identification of *Campylobacter* hotspots in areas with high human-livestock cohabitation, with chickens and goats acting as central transmission nodes, underscores the urgent need for targeted biosecurity measures, hygiene education, improved animal husbandry, and enhanced access to clean water and sanitation.

### Understanding One Health Disease Dynamics and Predicting Future Outbreaks

Identifying key variables that influence pathogen reservoirs and spillover events at high-risk interfaces and closely monitoring them (variables), researchers can detect early warning signals that indicate the potential emergence of zoonotic diseases, thereby improving our ability to predict future outbreaks and their transmission patterns. The integration of advanced machine learning techniques has revolutionized the field of disease surveillance. Both supervised and unsupervised models are increasingly being used to analyze vast datasets, revealing patterns and trends associated with emerging and re-emerging zoonotic diseases, including those linked to antimicrobial resistance (AMR). These models can process complex ecological, genetic, and epidemiological data to identify hidden correlations and predict disease emergence with greater accuracy. By leveraging these computational tools, researchers and public health authorities can develop more effective early warning systems, enhance preparedness strategies, and implement targeted interventions. This represents a significant advancement in global health security, ultimately strengthening efforts to prevent future outbreaks and mitigate their impact on human and animal populations.

### Policy and Practical Implications

This study highlighted key policy and practical implications for zoonotic disease prevention in Nepal, particularly in Bardia National Park’s buffer zones. A One Health approach is essential, integrating public health, veterinary, and conservation efforts into a national surveillance program. Water safety improvements, livestock management, and public awareness campaigns should address risk factors like water contamination and unsafe food practices. Conservation policies must consider zoonotic risks, ensuring wildlife protection does not increase human exposure. Strengthening laboratory capacity, early warning systems, and proactive health strategies will enhance disease prevention and public health resilience in Nepal. As we prepare this manuscript, efforts are underway to develop Nepal’s first Wildlife Health Surveillance Standard Operating Protocol (SOP).

### Study strengths and limitations

This study was the first of its kind to investigate the wildlife-human interface in Nepal, integrating Knowledge, Attitudes, and Practices (KAP) assessments with laboratory analyses. It establishes a foundational understanding of pathogen and disease ecology in a high-risk zoonotic interface.

Proactive surveillance of zoonotic pathogens demands significant resources, including technical expertise. While targeted pathogen detection provides valuable insights, it may not fully capture the complexity of disease ecology. Effective proactive surveillance must integrate diverse datasets, such as clinical findings and large-scale behavioral surveys, to provide a more comprehensive picture.

As a pilot study, this research does not incorporate these additional datasets, including a greater representation of wildlife taxa with high risk for zoonoses such as birds, bats and rodents. However, implementing a robust surveillance program will require a holistic approach that combines and analyzes critical variables influencing pathogen reservoirs, spillover events, and transmission dynamics.

## Conclusion

Proactive surveillance is essential for preventing zoonotic spillover by enabling early detection, targeted intervention, and risk mitigation. Given the critical role of wildlife in disease ecology, a One Health approach is crucial in vulnerable communities where human-livestock-wildlife interactions facilitate pathogen transmission. Routine pathogen screening in livestock, wildlife, and environmental samples allows for early identification of emerging threats like *Campylobacter*, *Salmonella*, and *Influenza A*, enabling timely interventions. Network analysis and spatial mapping help pinpoint high-risk areas, allowing for targeted biosecurity measures such as improved waste management, vaccination programs, and farm hygiene practices. Community-led disease monitoring enhances early warning systems, while environmental surveillance of contaminated water sources and shared grazing areas helps track indirect transmission pathways. Strengthening multisectoral collaboration between veterinarians, epidemiologists, ecologists, and public health officials improves data sharing and outbreak response. Investing in proactive surveillance also reduces economic burdens by preventing livestock losses, medical costs, and productivity declines. By integrating surveillance within a One Health framework, zoonotic spillover risks can be mitigated effectively, safeguarding both public health and economic resilience in at-risk communities.

## Data Availability

All data produced in the present work are contained in the manuscript

https://www.ncbi.nlm.nih.gov/bioproject/?term=PRJNA1225206

## Acknowledgement

This study was partially funded by the USAID Nepal Biodiversity Project, and we extend our gratitude to USAID for their generous support. We also wish to thank the Nepal Health Research Council (NHRC) for their guidance, support, and permission to conduct this study. The successful completion of this work would not have been possible without the support of the people of Thakurdwara Municipality and Bardia National Park. We appreciate the dedicated efforts of the field and laboratory teams from the CMDN and Intrepid Nepal Pvt. Ltd. We utilized AI tools to improve language in this manuscript.

## Data Availability

All the data are included in the manuscript and supplementary information. All the sequences obtained in this study have been uploaded to the NCBI GenBank under Bioproject-PRJNA1225206 with individual accession numbers within the Bioproject and are publicly available.

---

**Supplementary Table 1:** Selected Pathogens and Justification for including them in the study.

| **Pathogen** | **Selection Justification** |
| --- | --- |
| **Influenza A viruses** (e.g., H1N1) | Possess zoonotic potential, often crossing species barriers via intermediate hosts like poultry and pigs. Wild waterfowl serve as natural reservoirs, and the mixing of strains in animals can lead to new pandemic strains. Zoonotic transmission typically occurs through close contact with infected animals or contaminated environment [25,26]. |
| **Rhabdovirus** | The rabies virus exemplifies zoonotic transmission, residing primarily in wildlife like bats, raccoons, and skunks. Humans and domestic animals can contract the virus through bites or scratches from infected animals [27]. |
| **Paramyxovirus** | Though primarily transmitted between humans, some paramyxoviruses (e.g., measles and mumps) have zoonotic potential, particularly through contact with infected non-human primates. These viruses, often linked to respiratory infections, underscore the need for vigilant surveillance [28]. |
| **Filovirus** | Filoviruses, such as Ebola, have zoonotic origins, with fruit bats as natural reservoirs. Humans contract Ebola through contact with infected bats or by consuming bushmeat. Person-to-person transmission is also possible, requiring strict infection control during outbreaks [29,30]. |
| **Flavivirus** | Flaviviruses like Zika and West Nile are transmitted zoonotically via mosquito bites. While humans are typically dead-end hosts, zoonotic spillover can lead to localized outbreaks, raising public health concerns [31]. |
| **Coronavirus** | Coronaviruses, notably SARS-CoV-2, are known for their zoonotic potential, often originating in animals such as bats. Transmission occurs through close contact with infected animals or consumption of contaminated products, highlighting the need for proactive surveillance to prevent future pandemics [32,33]. |
| **Salmonella** | Frequently zoonotic, Salmonella infections in humans arise from contaminated animal products (e.g., raw eggs, poultry) or direct contact with infected animals, especially reptiles. Proper food handling and hygiene are crucial for prevention [34,35]. |
| **Brucella** | Primarily affecting livestock, Brucella bacteria transmit to humans via contact, ingestion of unpasteurized dairy, or handling contaminated tissues. Brucellosis presents flu-like symptoms, stressing the importance of pasteurization and livestock disease control [36,37]. |
| **Campylobacter** | Mainly originating in livestock, particularly poultry, Campylobacter can infect humans through undercooked meat, contaminated water, or direct contact with infected animals. It is a leading cause of bacterial gastroenteritis [38,39]. |
| **Mycobacteria** | Mycobacterium tuberculosis (TB) can infect various animals. Though human-to-human transmission is more common, zoonotic cases occur through contact with infected livestock, emphasizing the need for TB control in both human and animal populations [40,41]. |

**Supplementary Table 2:**
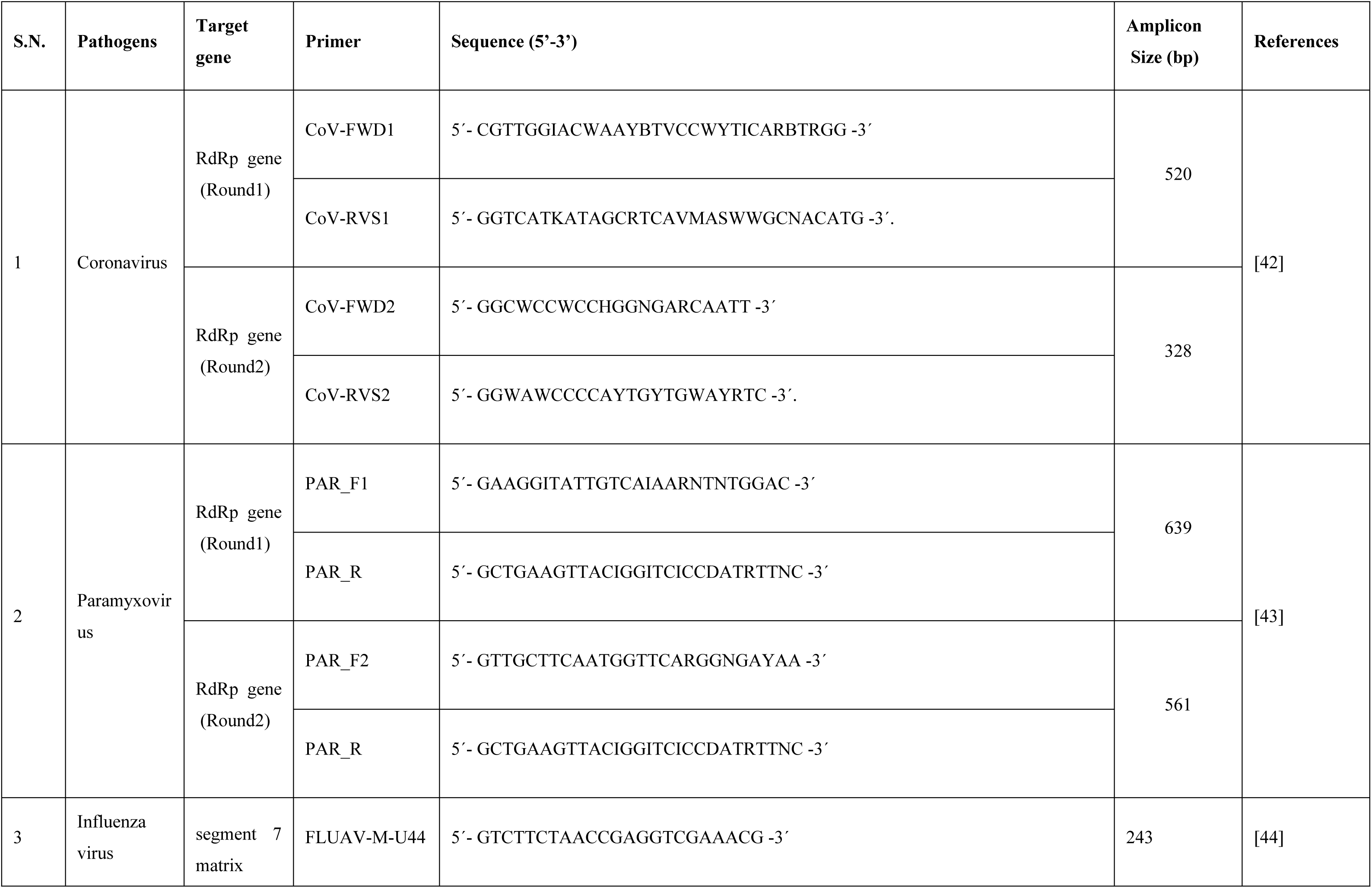
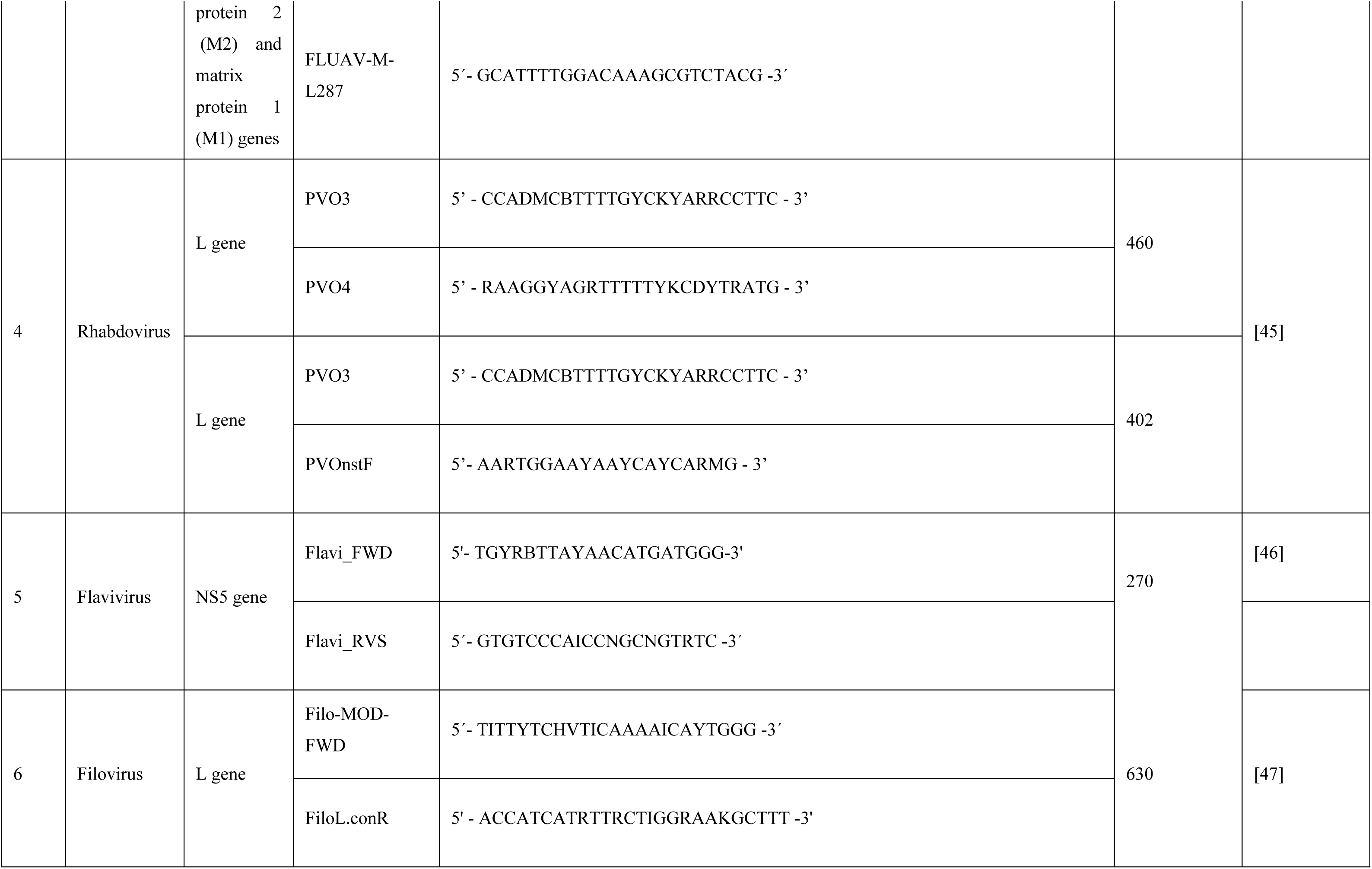
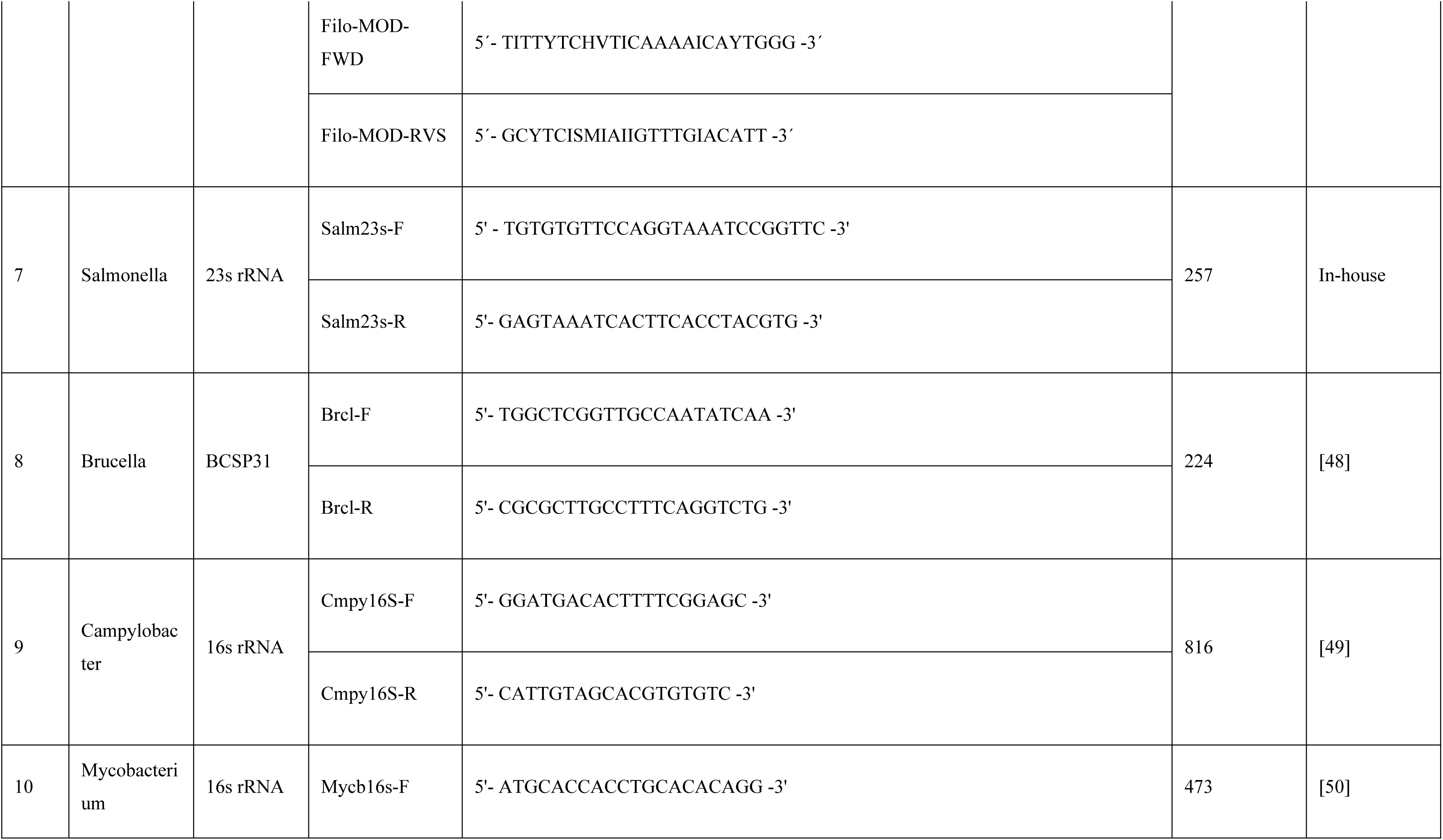
Pathogens with their target gene, PCR primer name and sequence, amplicon size, and references.

**Supplementary Table 3:** Detected *Campylobacteria* strains and their clinical significance.

| Abbreviation | Species Name | Clinical Significance |
| --- | --- | --- |
| **Cjeje** | *Campylobacter jejuni* subsp. *jejuni* | Most common cause of bacterial gastroenteritis worldwide.<br>Associated with **Guillain-Barré syndrome**, reactive arthritis.<br>Zoonotic; transmitted via poultry, milk, water [51–53]. |
| **Ccol** | *Campylobacter coli* | Second most common cause of **Campylobacter enteritis**.<br>Similar to *C. jejuni*, but slightly less prevalent.<br>Also zoonotic (pork, poultry, water) [53–55]. |
| **Cjedo** | *Campylobacter jejuni* subsp. *doylei* | Rare.<br>Associated with **bacteremia** and **gastroenteritis**, mostly in immunocompromised hosts [56]. |
| **Cla** | *Campylobacter lari* | Occasionally causes **gastroenteritis**.<br>Found in seagulls and marine environments.<br>Rarely causes **bacteremia** [57,58]. |
| **Cup** | *Campylobacter upsaliensis* | Emerging cause of **gastroenteritis**.<br>Zoonotic, commonly from dogs and cats [59–61]. |
| **Che** | *Campylobacter helveticus* | Rare human pathogen.<br>Isolated from domestic animals (dogs, cats) [53,59]. |
| **Cin** | *Campylobacter insulaenigrae* | Rare, isolated from marine mammals.<br>Minimal known clinical significance in humans [62–65]. |
| **Ccl** | *Campylobacter species clone* | Broad term for genetically related but undefined species.<br>Significance varies depending on the clone [66,67]. |
| **Chyla** | *Campylobacter hyointestinalis* subsp. *lawsonii* | Primarily associated with pigs.<br>Rare human cases of **gastroenteritis** [68,69]. |
| **Chyhy** | *Campylobacter hyointestinalis* subsp. *hyointestinalis* | Similar to *C. hyointestinalis* subsp. *lawsonii*.<br>Rare human infections [70–73]. |
| **Cfefe** | *Campylobacter fetus* subsp. *fetus* | Causes **bacteremia**, **septic abortion**, and systemic infections, especially in immunocompromised people.<br>Zoonotic (sheep, cattle) [74,75]. |
| **Cfeve** | *Campylobacter fetus* subsp. *venerealis* | Primarily veterinary significance (bovine genital campylobacteriosis).<br>Rare human infections [76–78]. |
| **Cmu** | *Campylobacter mucosalis* | Rarely isolated from humans.<br>Possible links to **intestinal diseases** in animals [79,80]. |
| **Csp** | *Campylobacter sputorum* | Mostly found in cattle and sheep.<br>Human infections are exceedingly rare [81,82]. |
| **Cho** | *Campylobacter hominis* | ▪ Rare human isolate.<br>▪ Found in fecal samples; pathogenic potential unclear [83,84]. |
| **Cgr** | *Campylobacter gracilis* | ▪ Found in the **oral cavity**.<br>▪ Associated with **periodontal disease** and **oral abscesses** [85,86]. |
| **Cre** | *Campylobacter rectus* | ▪ Strongly associated with **periodontal disease**.<br>▪ May contribute to **systemic diseases** through oral infections [87,88]. |
| **Csh** | *Campylobacter showae* | ▪ Oral flora.<br>▪ Potentially involved in **periodontal disease** [89,90]. |
| **Ccon** | *Campylobacter concisus* | ▪ Emerging pathogen in **gastrointestinal diseases**, including **Crohn's disease** and **ulcerative colitis**.<br>▪ Found in the oral cavity and gut [91,92]. |
| **Ccu** | *Campylobacter curvus* | ▪ Part of oral flora.<br>▪ Associated with **periodontal disease** [93,94]. |
| **Clan** | *Campylobacter lanienae* | ▪ Found in humans and animals.<br>▪ Possible gastrointestinal pathogen; limited data [95–97]. |

